# EFFICACY OF THE MEASLES-MUMPS-RUBELLA (MMR) VACCINE IN THE REDUCING THE SEVERITY OF COVID-19: AN INTERIM ANALYSIS OF A RANDOMISED CONTROLLED CLINICAL TRIAL

**DOI:** 10.1101/2021.09.14.21263598

**Authors:** Edison Natal Fedrizzi, Juliana Balbinot Reis Girondi, Thiago Mamoru Sakae, Sérgio Murilo Steffens, Aldanéa Norma de Souza Silvestrin, Grace Serafim Claro, Hugo Alejandro Iskenderian, Bianca Hillmann, Liliam Gervasi, Alberto Trapani, Patricia de Amorim Rodrigues, Amanda de Souza Vieira, Scheila Monteiro Evaristo, Francisco Reis Tristão, Fabiano da Silva Muniz, Maria Veronica Nunes, Nicole Zazula Beatriz, Jhonathan Elpo, Amanda Tiedje, Louise Staudt Siqueira, Marilin Sens, Vitor Nandi, Jessica Goedert Pereira, Gustavo Costa Henrique, Ana Paula Fritzen de Carvalho, Ramon Carlos Pedroso de Morais, on behalf of the MMRCoV Study Group

**Author notes:** **CORRESPONDIG AUTHOR** Edison Natal Fedrizzi, Clinical Research Center of University Hospital, Federal University of Santa Catarina, Florianópolis, Brazil, Av Governador José Boabaid, 272, Zip Code: 88·037-200, Florianópolis, Santa Catarina state, Brazil.

## Abstract

**Background:** COVID-19 is still a challenge, both with regard to its treatment and to the actual efficacy of the vaccines available to date, especially with the emergence of new variants. We evaluated the efficacy of the measles-mumps-rubella (MMR) vaccine in preventing SARS-CoV-2 infection and severity of COVID-19 in health workers.

**Methods:** This analysis includes data from one ongoing blinded, randomized, placebo-controlled trial with participants aged 18-60 years were randomly assigned to receive the MMR vaccine or a placebo. The primary efficacy analysis included all participants with a positive nasopharyngeal RT-PCR test since their inclusion.

**Results:** The MMR vaccine did not prevent the SARS-CoV-2 infection. Participants in the MMR group, compared with those in the placebo group, had a 48% risk reduction in symptomatic COVID-19 (RR = 0.52; 95% CI: 0.33–0.83; p=0.004) and a 76% risk reduction in COVID-19 treatment (RR = 0.24; 95% CI: 0.06 – 0.88; p = 0.020) with one dose and a 51% risk reduction in COVID-19 symptoms (RR = 0.49; 95% CI: 0.31 – 0.78; p = 0.001) and a 78% risk reduction in COVID-19 treatment (RR = 0.22; 95% CI: 0.06 – 0.82; p = 0.015) with two doses.

**Conclusions:** This interim analysis of an ongoing clinical trial suggests that compared with a placebo, the vaccine reduces the risk of COVID-19 symptoms and reduces the need for COVID-19 treatment.

**Clinical Trials Registry:** Brazilian Clinical Trials Registry (ReBEC n° RBR-2xd6dkj - https://ensaiosclinicos.gov.br/rg/RBR-2xd6dkj).

**HIGHLIGHTS:**

1. The MMR vaccine can stimulate the innate immunity inducing a nonspecific protection against other infections, called heterologous immunity.
2. Repeated exposure to the antigen (innate immune response training) results in an extension of the action time of this immune response (innate immune response memory) and consequently in protection against other infections (heterologous immunity) for a longer time.
3. The MMR vaccine has been used by national immunization programs in the world for many years, it is very safe and can be stored and distributed at 2-8°C, making it particularly suitable for global distribution.
4. Among participants who received at least one dose, compared with those in the placebo group, participants in the MMR group had a significant risk reduction in symptomatic COVID-19 and of cases requiring treatment.
5. The use of MMR vaccine can be useful in several populations in the world that do not have access to the COVID-19 vaccine and in a future epidemic or pandemic as an emergency measure until specific treatments or vaccines for each case are available to the general population.

## 1. Introduction

In December 2019, China began to observe cases of severe acute respiratory syndrome caused by a novel coronavirus (severe acute respiratory syndrome coronavirus 2 - SARS-CoV-2), later named coronavirus disease 2019 (COVID-19) [1]. This infection spread rapidly around the world and was declared a pandemic by the World Health Organization (WHO) on March 11, 2020, the first pandemic caused by a coronavirus [2].

Recent studies have shown that possibility the greater the amount of virus inhaled by an individual, the higher the viral load and perhaps the greater the severity of the disease. SARS-CoV-2 has the ability to inhibit the immune response, especially the production of interferons, which may facilitate this increase in the viral load [3-6]. The most effective way to reduce severe cases and deaths is through the prevention of SARS-CoV-2 infection using a vaccine. However, although vaccines are already being administered in several countries, vaccination of the entire indicated population, especially in less developed countries, is not expected in the next few years.

Some studies have raised the possibility of using some vaccines to improve the immune response, such as BCG, measles or triple viral (measles, mumps and rubella), oral poliomyelitis or other vaccines, preferably with live attenuated antigen, due to their robust and long-lasting immune response [7-9]. This nonspecific immune response produces a potent protective barrier that can prevent cell invasion by another virus [10,11].

The objective of using a vaccine with live and attenuated microorganisms is to stimulate the innate immune response; repeated exposure to the antigen (innate immune response training) results in an extension of the action time of this immune response (innate immune response memory) and consequently in protection against other infections (heterologous immunity) for a longer time [7,8,10-15]. Even if this effect lasts for a limited period of time, it can greatly contribute to reducing the spread of infection in the early stages of a pandemic and/or reducing the severity of the disease, thus preventing hospitalizations and deaths [10,12,16-18]. Therefore, if one of these vaccines does not completely prevent infection, it could reduce the inhaled viral load and reduce the severity of COVID-19. This possible heterologous immunity of the measles-mumps-rubella (MMR) vaccine may be associated with potent stimulation of innate immunity, leading to a decrease in viral load via cellular immunity [10,13,14,16] and possible humoral immunity due to some similarities between the glycoproteins of the measles and rubella viruses and SARS-CoV-2 [9,19] and the action of antibodies against mumps [20].

The objective of this study was to evaluate whether the MMR vaccine, compared with a placebo, protects against COVID-19 or at least decreases the symptoms and severity of this infection in health workers.

## 2. Methods

### 2.1. Study design and participants

This is a randomized, double-blind, placebo-controlled, phase 3, clinical trial to evaluate efficacy of the MMR vaccine against SARS-CoV-2 induced COVID-19 in health workers from Florianópolis, Brazil, seen at the University Hospital at Federal University of Santa Catarina. The trial protocol was reviewed and approved by Ethics Committees of Federal University of Santa Catarina (n° 4.254.143) and National Research Ethics Committee (n° 4.274.984).

All study volunteers met all of the following criteria to participate in the study (inclusion criteria): 1) male and female health workers aged 18 to 60 years (inclusive) at the first visit (V1); 2) volunteers who, in the opinion of the researchers, were able to meet the requirements of the study protocol. The following exclusion criteria were used to select volunteers at V1: 1) women who were pregnant or wishing to become pregnant during the study period; 2) women who were breastfeeding; 3) individuals with immune system deficiency or receiving or had received treatments such as chemotherapy, radiotherapy, high corticosteroid doses (>20 mg/day for ≥14 days) or immunosuppressants in the past 90 days; 4) individuals who received immunoglobulins or blood products in the past 180 days; 5) patients with decompensated autoimmune diseases, such as systemic lupus erythematosus, multiple sclerosis, Guillain-Barré syndrome, myasthenia gravis, scleroderma; 6) individuals who received any attenuated vaccine in the last 30 days or inactivated vaccine in the last 15 days; 7) individuals with a fever >37.8°C and/or inflammatory/infectious process in the oropharynx; 8) individuals with a history of a positive test for SARS-CoV-2 by RT-PCR (reverse transcription polymerase chain reaction) or rapid test or serology; 9) individuals with a clinical history and/or physical examination indicating COVID-19, such as fever (>38°C), painful swallowing, cough, laboured breathing, loss of taste and loss of smell; 10) individuals with anaphylactic-type hypersensitivity to some vaccine component (especially neomycin); 11) individuals with any other clinical condition that, at the discretion of the investigator, could interfere in the evaluation of the study results or bring some risk to the volunteer; and 12) individuals who were participating or intended to participate in another study evaluating the use of another vaccine. All participants gave written informed consent.

### 2.2. Randomisation and blinding

A total of 430 health workers were included in the study. The participants selected for the study were mainly COVID-19 frontline health workers, of both sexes, aged between 18 and 60 years. The participants were divided into 2 groups randomly allocated at a 1.5:1 ratio, with the MMR group consisting of 246 volunteers and the placebo group (saline solution) consisting of 178 volunteers. Randomisation list was prepared by the study statistician (TMS) and washeld by the unmasked study nurse who prepared the vaccines for administration, with all other trial staff masked to group allocation. The vaccine and placebo were prepared out of sight of the participants and all study staff and syringes were covered with an opaque material until ready for administration.

### 2.3. Procedures

Those participants with a negative rapid test underwent sample collection from the nasopharynx for SARS-CoV-2 RT-PCR testing and received the appropriate injection (vaccine or placebo); the same occurred at V3 (after 60 days), when the participants received the second injection. The volunteers received two doses, at an interval of 8 weeks of 0.5 mL of subcutaneous MMR vaccine or placebo. The MMR vaccine used in this study was manufactured by Fiocruz - Biomanguinhos Laboratory (Rio de Janeiro, Brazil) and contains three live attenuated viruses that are highly immunogenic, providing lasting protection for most vaccinated individuals after two doses [21,22]. It was licenced for commercialization in the 1970s and included in vaccination schedules in the Americas and Europe at the same time [22]. The MMR vaccine is highly safe and minimally reactive, has minimal side effects and is widely used around the world [15]. Adverse reactions are rarely observed after its administration [21,22].

Adverse events were recorded by the participants for 30 consecutive days after each vaccination. In the subsequent visits, every 30 days (until V7), a new clinical evaluation, rapid test and RT-PCR were performed regardless of whether the participant had any symptoms suggestive of COVID-19. When COVID-19 was suspected, the participant made an extra visit to the Research Center.

The study lasted approximately six months after V1, with 7 visits at 4-week intervals.

A case was defined as a volunteer with an RT-PCR test positive for SARS-CoV-2, regardless of whether the individual was symptomatic. To assess severity, COVID-19 was classified into four groups: 1) asymptomatic – positive RT-PCR and no symptoms; 2) mild – positive RT-PCR and mild respiratory or clinical symptoms, with home monitoring; 3) moderate – positive RT-PCR and respiratory or clinical symptoms that required treatment such as anticoagulation, corticosteroid and antibiotic therapy or hospitalization; and 4) severe: positive RT-PCR with respiratory symptoms or other complications that required admission to the intensive care unit or resulted in death [23,24].

### 2.4. Outcomes

The first primary endpoint was the efficacy of the MMR vaccine against symptomatic COVID-19 confirmed in participants with symptoms (at least one symptom, such as fever, cough, laboured breathing, chills, painful swallowing, muscle ache, loss of smell, loss of taste, diarrhoea or vomiting) and a positive RT-PCR test. The secondary endpoints were preventing SARS-CoV-2 infection, as confirmed by RT-PCR, in participants with or without symptoms and the efficacy of the MMR vaccine against moderate and severe COVID-19.

We also evaluated whether there was a significant difference between the groups with or without COVID-19 regarding sex, age, race, profession, adverse events and associated comorbidities.

### 2.5. Statistical analysis

Assuming an expected infection rate of 20% in the group not exposed to vaccination and 10% in the vaccinated group, with an alpha error of 5%, power of 80%, and group ratio of 1.5:1, the calculated minimum sample size required was 405 volunteers. Adding a risk of unexpected losses of 5%, the minimum sample size required for this study was 420 volunteers. Our study included 430 volunteers.

The efficacy of the MMR vaccine compared to placebo in the prevention of COVID-19 was demonstrated by calculating vaccine efficacy as (1-RR) * 100, where the relative risk (RR) was defined as the proportion of the rate of individuals with COVID-19 in group 1 (MMR vaccine) over the rate of individuals with COVID-19 in group 2 (control).

In the Intention-to-treat population analysis (ITT), all randomised study participants were included. The observation time started when the first dose of the vaccine/placebo was administered. Model based multiple imputation (MI) was used for both primary and secondary outcomes.

A Per-Protocol analysis (PP) was performed with all randomised study participants completing the whole study period (complete cases). For a specific analysis, study participants with missing data on any of the variables in the model was excluded from the analysis.

The reduction in COVID-19 severity was evaluated by comparing cases of asymptomatic, mild, moderate and severe disease between the group that received the MMR vaccine and the group that received placebo.

## 3. RESULTS

In the period from 29 July 2020 to 20 February 2021, a total of 430 health workers aged 18 to 60 years were selected for the study. Of these, six were excluded for having a positive rapid test for SARS-CoV-A total of 424 participants were randomly allocated to receive the MMR vaccine or placebo. A total of 246 volunteers from the vaccine group and 178 from the placebo group received at least one dose. All of these participants were included in the efficacy analysis, with a median follow-up of five months (Fig. 1). Of these 424 participants, 76% were women, and 89% were white. The mean age was 38 years, with 15% being over 50 years old. Among the health workers, 23% were doctors, 23% were health technicians and 21% were nurses. The most frequent comorbidities among the participants were hypertension (9.4%) (Table 1). There were six cases (5.8%) of probable reinfection, all with positive RT-PCR tests at an interval of more than three months, interspersed with negative tests. All participants were asymptomatic or had mild symptoms. It was not possible to perform genetic sequencing. Another eight participants (7.8%) had positive RT-PCR results for two or three months (Table S1).

**Figure 1.**
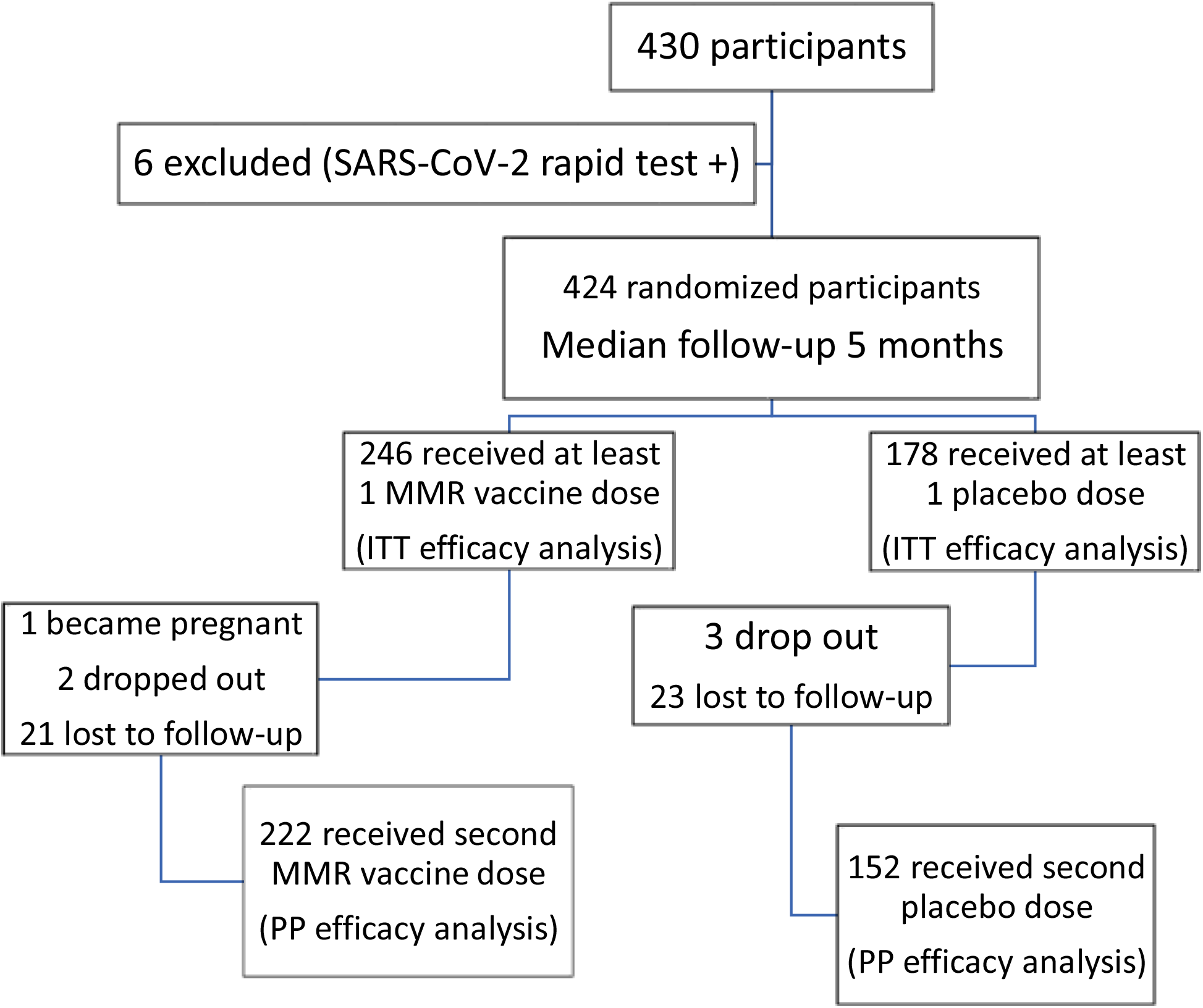
Inclusion and randomization of participants in the MMRCoV study. The diagram represents all participants included in the study from 29 July 2020 to 20 February 2021; 424 participants were included in the efficacy analysis, with a median follow-up of 5 months, ranging from 3 to 7 months. The efficacy analysis began at the first application of the medication (vaccine or placebo).

**Table 1.** Sociodemographic and clinical characteristics of the participants of the MMRCoV study.

| Characteristic | MMR Group | Placebo Group | Total | p-value |
| --- | --- | --- | --- | --- |
| **Sex** |  |  |  | 0.491 |
| male | 56 (54.9) | 46 (45.1) | 102 (24.05) |  |
| female | 190 (59.0) | 32 (41.0) | 322 (75.95) |  |
| **Age** |  |  |  | 0.436 |
| mean (± standard deviation) | 37.49 (10.53) | 38.30 (10.61) | | |
| **Age group** |  |  |  | 0.919 |
| 18-30 | 76 (30.9) | 51 (28.7) | 127 (30.0) |  |
| 31-40 | 73 (29.7) | 51 (28.7) | 124 (29.2) |  |
| 41-50 | 61 (24.8) | 47 (26.4) | 108 (25.5) |  |
| 51-60 | 36 (14.6) | 29 (16.3) | 65 (15.3) |  |
| **Race** |  |  |  | 0.630 |
| White | 220 (89.4) | 158 (88.8) | 378 (89.2) |  |
| Black | 5 (2.0) | 6 (3.4) | 11 (2.6) |  |
| Mixed | 19 (7.7) | 11 (6.2) | 30 (7.1) |  |
| Asian | 2 (0.8) | 3 (1.7) | 5 (1.2) |  |
| **Occupation** |  |  |  | 0.354 |
| doctor | 48 (19.5) | 51 (28.8) | 99 (23.4) |  |
| nurse | 54 (22.0) | 38 (21.5) | 92 (21.7) |  |
| psychologist | 7 (2.8) | 4 (2.3) | 11 (2.6) |  |
| pharmacist/biomedical professional | 11 (4.5) | 2 (1.1) | 13 (3.1) |  |
| physical therapist | 3 (1.2) | 9 (5.1) | 12 (2.8) |  |
| health technician* | 64 (26.0) | 35 (19.8) | 99 (23.4) |  |
| social worker | 4 (1.6) | 1 (0.6) | 5 (0.7) |  |
| **nutritionist** | 5 (2.0) | 2 (1.1) | 7 (1.7) |  |
| **dentist** | 6 (2.4) | 6 (1.4) | 12 (2.8) |  |
| **health student** | 21 (8.5) | 9 (5.1) | 30 (7.1) |  |
| **other<sup>†</sup>** | 23 (9.3) | 20 (11.3) | 43 (10.2) |  |
| **Comorbidities** |  |  |  |  |
| **hypertension** | 19 (7.7) | 21 (11.8) | 40 (9.4) | 0.179 |
| **diabetes** | 5 (2.0) | 4 (2.2) | 9 (2.1) | 1.000 |
| **pneumopathy** | 17 (6.9) | 11 (6.2) | 28 (6.6) | 0.845 |
| **obesity** | 14 (5.7) | 14 (7.9) | 28 (6.6) | 0.430 |
| **immunosuppression** | 0 (0.0) | 0 (0.0) | 0 (0.0) | 1.000 |
| **hypothyroidism** | 12 (4.9) | 11 (6.2) | 23 (5.4) | 0.665 |
| **smoking** | 18 (7.3) | 7 (3.9) | 25 (5.9) | 0.209 |

\*Health technicians included nursing, radiology, clinical analysis, cytopathology and laboratory technicians.
†Other professionals included in the study were health agents, biomedical professionals, firefighters and surveillance services and hospital care workers.

There was no significant difference in sex, race, age or profession when evaluating the cases of COVID-19. When assessing comorbidities, hypertension, obesity and pneumopathy were the most frequent in both groups. However, we observed a higher risk of participants with diabetes mellitus becoming infected with SARS-CoV-2 (RR = 11.04; 95% CI: 2.33-52.35; p = 0.0009) (Table 2).

**Table 2.**
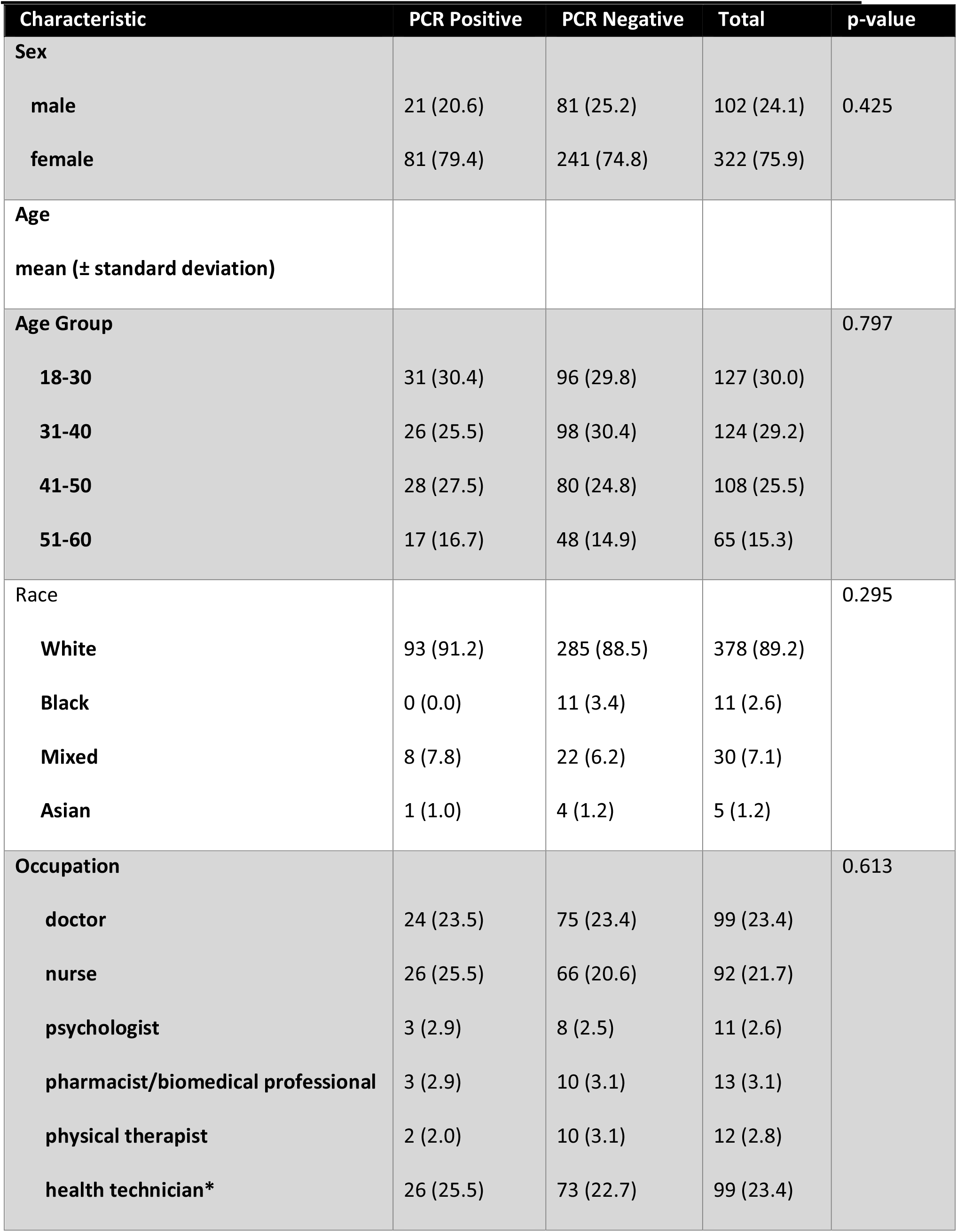
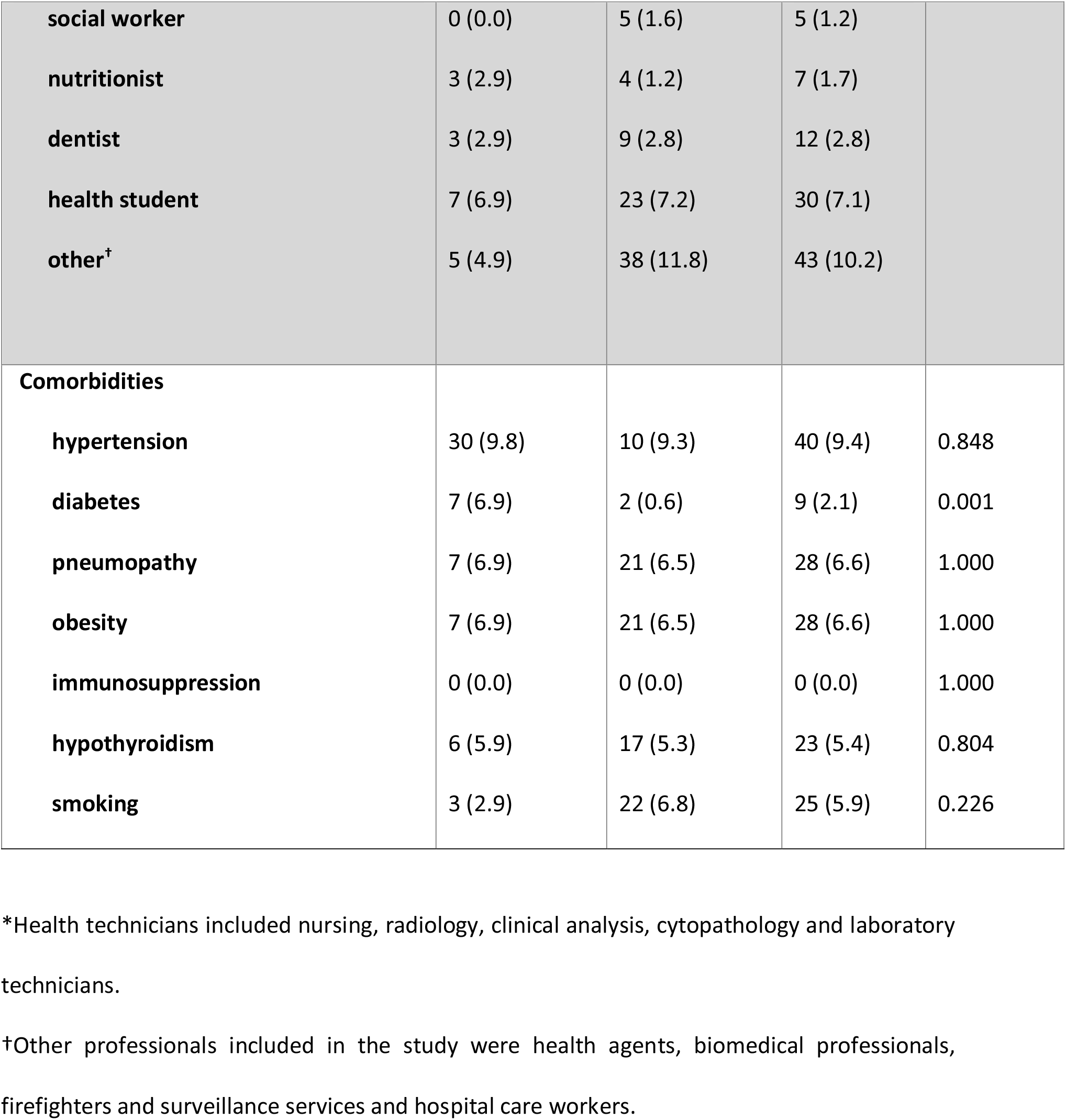
Sociodemographic and clinical characteristics based on PCR results for MMRCoV study participants.

| Characteristic | PCR Positive | PCR Negative | Total | p-value |
| --- | --- | --- | --- | --- |
| **Sex** |  |  |  |  |
| male | 21 (20.6) | 81 (25.2) | 102 (24.1) | 0.425 |
| female | 81 (79.4) | 241 (74.8) | 322 (75.9) |  |
| **Age** |  |  |  |  |
| mean (± standard deviation) | | | | |
| **Age Group** |  |  |  | 0.797 |
| 18-30 | 31 (30.4) | 96 (29.8) | 127 (30.0) |  |
| 31-40 | 26 (25.5) | 98 (30.4) | 124 (29.2) |  |
| 41-50 | 28 (27.5) | 80 (24.8) | 108 (25.5) |  |
| 51-60 | 17 (16.7) | 48 (14.9) | 65 (15.3) |  |
| **Race** |  |  |  | 0.295 |
| White | 93 (91.2) | 285 (88.5) | 378 (89.2) |  |
| Black | 0 (0.0) | 11 (3.4) | 11 (2.6) |  |
| Mixed | 8 (7.8) | 22 (6.2) | 30 (7.1) |  |
| Asian | 1 (1.0) | 4 (1.2) | 5 (1.2) |  |
| **Occupation** |  |  |  | 0.613 |
| doctor | 24 (23.5) | 75 (23.4) | 99 (23.4) |  |
| nurse | 26 (25.5) | 66 (20.6) | 92 (21.7) |  |
| psychologist | 3 (2.9) | 8 (2.5) | 11 (2.6) |  |
| pharmacist/biomedical professional | 3 (2.9) | 10 (3.1) | 13 (3.1) |  |
| physical therapist | 2 (2.0) | 10 (3.1) | 12 (2.8) |  |
| health technician* | 26 (25.5) | 73 (22.7) | 99 (23.4) |  |
| **social worker** | 0 (0.0) | 5 (1.6) | 5 (1.2) |  |
| **nutritionist** | 3 (2.9) | 4 (1.2) | 7 (1.7) |  |
| **dentist** | 3 (2.9) | 9 (2.8) | 12 (2.8) |  |
| **health student** | 7 (6.9) | 23 (7.2) | 30 (7.1) |  |
| **other<sup>†</sup>** | 5 (4.9) | 38 (11.8) | 43 (10.2) |  |
| **Comorbidities** |  |  |  |  |
| **hypertension** | 30 (9.8) | 10 (9.3) | 40 (9.4) | 0.848 |
| **diabetes** | 7 (6.9) | 2 (0.6) | 9 (2.1) | 0.001 |
| **pneumopathy** | 7 (6.9) | 21 (6.5) | 28 (6.6) | 1.000 |
| **obesity** | 7 (6.9) | 21 (6.5) | 28 (6.6) | 1.000 |
| **immunosuppression** | 0 (0.0) | 0 (0.0) | 0 (0.0) | 1.000 |
| **hypothyroidism** | 6 (5.9) | 17 (5.3) | 23 (5.4) | 0.804 |
| **smoking** | 3 (2.9) | 22 (6.8) | 25 (5.9) | 0.226 |

\*Health technicians included nursing, radiology, clinical analysis, cytopathology and laboratory technicians.
†Other professionals included in the study were health agents, biomedical professionals, firefighters and surveillance services and hospital care workers.

Among the 424 participants who entered the study and received at least one dose of the MMR vaccine or placebo, there were 102 cases of COVID-19, with no significant difference in cases of infection between the groups (RR = 0.91; 95% CI: 0.65-1.28; p = 0.348) (Table 3).

**Table 3.**
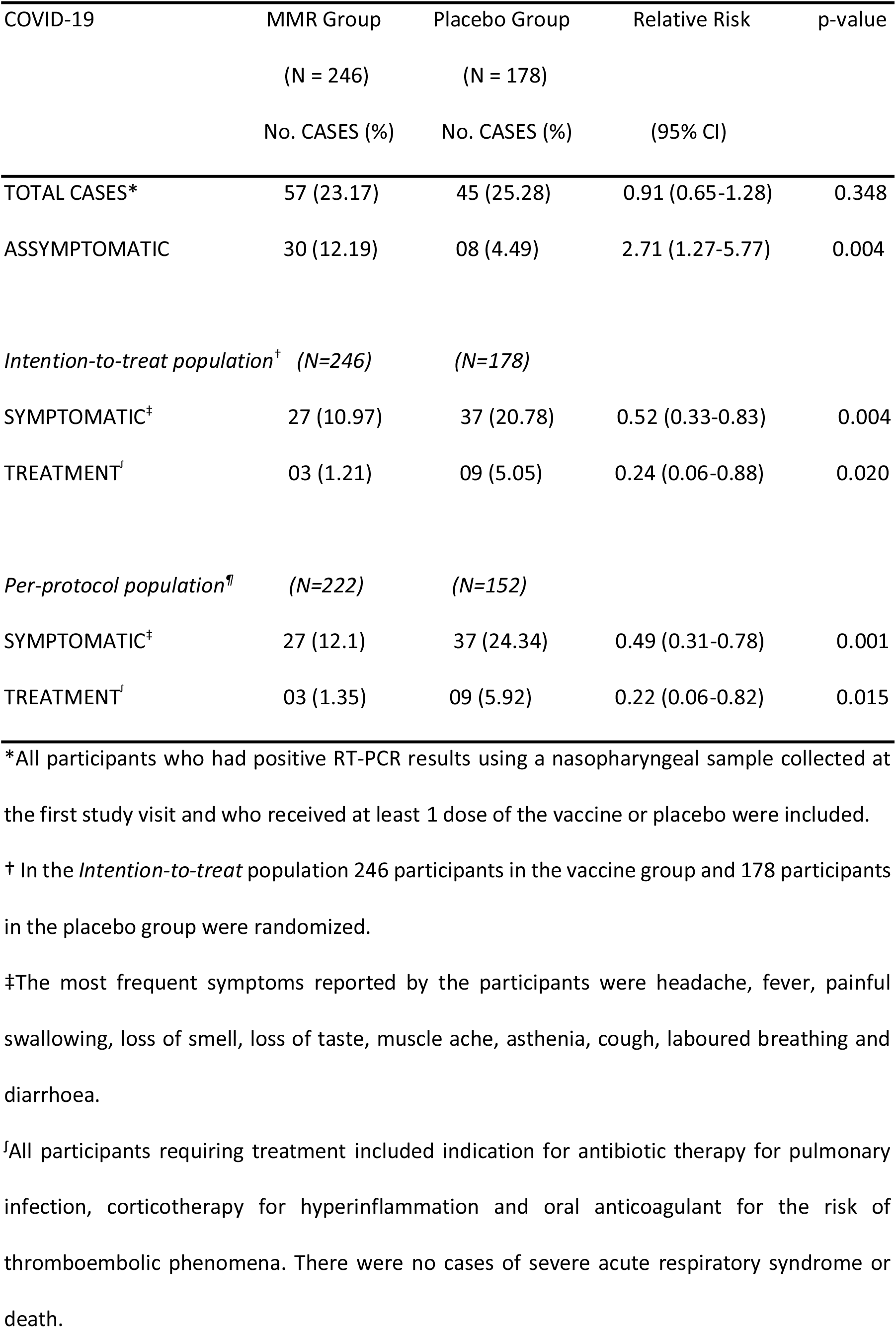
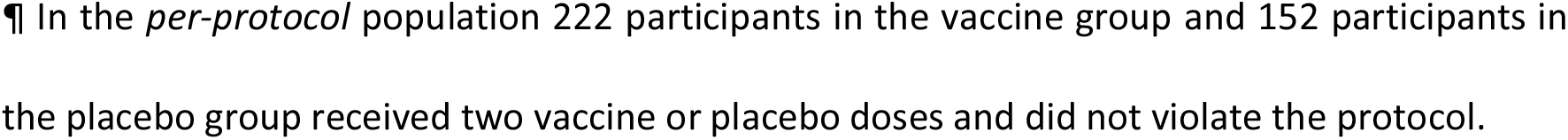
Relative risk of COVID-19 and treatment after one and two vaccine doses in the MMRCoV study.

Of the total number of RT-PCR-positive cases in each group, we observed a higher frequency of asymptomatic cases in the MMR group (12.19%) than in the placebo group (4.49%) (RR = 2.71; 95% CI: 1.27-5.77; p = 0.004). In the *intention-to-treat population* (ITT) analysis, where the participants received at least one dose of the vaccine (n = 246) or placebo (n = 178), the MMR group had a 48% risk reduction for symptomatic COVID-19 (RR = 0.52; 95% CI: 0.33-0.83; p = 0.004). Regarding the need for treatment for COVID-19 a 76% risk reduction was observed (RR = 0.24; 95% CI: 0.06-0.88; p = 0.020) in the vaccinated group. In the *per-protocol susceptible population* (PP) analysis, where the participants received both doses of the vaccine (n = 222) or placebo (n = 152) and had no deviation from the protocol, the MMR group had a 51% risk reduction for symptomatic COVID-19 (RR = 0.49; 95% CI: 0.31-0.78; p = 0.001). For the need for treatment for COVID-19 a 78% risk reduction was observed (RR = 0.22; 95% CI: 0.06-0.82; p = 0.015) (Table 3). There were no cases of severe COVID-19.

There were 33 cases where an asymptomatic participant with a positive PCR test received the vaccine or placebo (visit 1 and 3). Among the participants who received the vaccine, 69.6% remained asymptomatic (23/33), and among those who received placebo 30.3% (10/33) (p=NS).

The MMR vaccine was safe and well tolerated, with minimal adverse events, especially at the injection site. No serious adverse events, either local or systemic, were reported in the study in the vaccinated group or placebo group (Table S2).

## 4. DISCUSSION

The administration of at least one dose of the MMR vaccine resulted in a significant reduction of 48% in symptomatic COVID-19 and of 76% in the need for treatment. When the participant used two doses, the reduction was 51% and 78%, respectively. These results confirm several previous studies that observed a reduction of infections through heterologous immunity via stimulation of innate immunity by attenuated vaccines [7,13-15] and specifically by the MMR vaccine [10,13,14]. Although several studies in the literature suggest that the MMR vaccine has some protective action against COVID-19 [7,16,18-20], this is the first double-blind, placebo-controlled, randomized clinical trial that demonstrated this action of the MMR vaccine against COVID-19.

The percentage of positive cases in the study was 24% over a median follow-up period of 5 months. This high percentage is explained by the fact that the participants all work in the health sector and many work on the front line. Of the positive cases, 25.5% of nurses and 23.5% of doctors had COVID-19, diagnosed by RT-PCR. In a systematic review and meta-analysis of the prevalence of COVID-19 in health workers diagnosed by RT-PCR, was observed a high prevalence of infection in the health sector (11%), and nurses were also the most infected (48%), followed by doctors (43%) [17].

In the same way that specific vaccines for COVID-19 did not show efficacy in preventing asymptomatic infection [25-29], our study also did not show this efficacy. However, most cases of COVID-19 in the group that received the vaccine remained asymptomatic and did not require treatment or hospitalization, even for those with a positive RT-PCR test at the first study visit, suggesting rapid stimulation of innate immunity. These results corroborate the hypotheses of several studies that the triple viral vaccine has protective action with regard to disease severity but not disease prevention. Some explanations that indicate that the MMR vaccine may have some action against COVID-19 include 1) potent stimulation of the innate immune response and its memory [8,10-12]. Recent reports indicate that COVID-19 can suppress the innate immune response [5]. 2) The MMR viruses and the coronavirus are RNA viruses, and perhaps, this may further benefit the stimulation of the innate immune response using the MMR vaccine [7]. 3) A 30 amino acid sequence is homologous among the SARS-CoV-2 spike glycoprotein (PDB:6VSB), the measles virus fusion glycoproteins (PDB:5YW_B) and the rubella virus envelope (PDB:4ADG_A). These regions may be antigenic epitopes that stimulate humoral immunity, generating antibodies that could protect against COVID-19. This could be one of the explanations for the lower susceptibility of children to COVID-19 [9,19]. 4) The macrodomains of the SARS-CoV-2 virus and rubella virus have an amino acid sequence with 29% identity. Hypothetically, these SARS-CoV-2 macrodomains could be recognized by anti-rubella antibodies and provide a certain degree of protection. One study showed that the levels of antibodies against rubella (IgG) increased in patients with COVID-19, similar to cases of a second infection with rubella [19]. 5) There is significant evidence of an inverse correlation between the levels of antibodies against mumps and COVID-19 severity, i.e., the higher the concentration of anti-mumps IgG antibodies, the lower is the severity of COVID-19 [20].

The duration of protection of heterologous immunity is not yet well defined. In our study, the median follow-up was 5 months. Several studies have shown that the memory of innate immunity (stimulated by innate immunity training) may last for 1 year or more [16,30].

This study had some limitations. Although the number of participants was small, the median follow-up of five months was sufficient for a statistical analysis to demonstrate significance. A second issue to address is the duration of the action of the MMR vaccine in COVID-19 disease progression. We still do not know how long it lasts, but the study is still ongoing to try to answer this question. It would be extremely important to have evaluated the cellular immune response and viral load between the vaccinees and placebo, as this would be an indicator of the effectiveness of MMR vaccine in inducing innate immune responses that could reduce viral load. We were unable to conducted genetic sequencing of suspected cases of reinfection due to laboratory challenges. We did not have information on the dosages of antibodies in the individuals who developed COVID-19 and were vaccinate or not with MMR. It was not possible to perform the genotyping of Sars-CoV-2 in infected patients to assess the possible variants. The age range evaluated was 18-60 years, and we cannot guarantee that the observed results will be the same in people above and below these ages. The results of the trial were obtained from health workers, who are generally healthy and it is not possible to extrapolate these same results to the general population.

## 5. CONCLUSIONS

The results of this analysis showed that the MMR vaccine can significantly decrease the rate of symptomatic COVID-19 and of cases requiring treatment. As this result we can propose that the MMR vaccine would be useful in several populations in the world that do not have access to the COVID-19 vaccine and in a new epidemic or pandemic as an emergency measure until specific treatments or vaccines for each case are available to the general population. These results are intriguing but would need confirmation with a larger study enrolling a general population at higher risk of severe disease or death. However, importantly, the MMR vaccine does not replace specific vaccines against COVID-19.

## Data Availability

The results of this study are preliminary and the study is ongoing. A complete de-identified patient dataset will be available upon completion of clinical trials and publication of the results of the completed study upon request to the corresponding author. Proposals will be reviewed and approved by the MMRCoV Study group on the basis of scientific merit and absence of competing interests. Once the proposal has been approved, data will be transferred after the signing of a data access agreement and a confidentiality agreement.

## Acknowledgments

We would like to thank the Federal University of Santa Catarina (UFSC) and Santa Catarina State Research and Innovation Foundation (FAPESC) for financing the study; University Hospital Prof Polydoro Ernani de São Thiago – HU/UFSC/EBSERH for the research site where the study was carried out; Fiocruz-Biomanguinhos Laboratory for the supply of vaccines; Santa Catarina State Health Secretariat (SES/SC) and Central Public Health Laboratory (LACEN) for conducting the RT-PCR tests for SARS-CoV-2; Florianópolis Municipal Health Department (SMS/PMF), Health Center of São José Citty Hall (SMS/PMSJ) and UFSC Health Sciences Center (CCS/UFSC) for the supply of consumables for the study, and all volunteers who participated in this study.

## Author contributions

Edison Natal Fedrizzi: Conceptualization, Methodology, Writing original draft preparation, Project administration; Juliana Balbinot Reis Girondi: Supervision, Investigation; Thiago Mamoru Sakae: Visualization, Investigation, Data curation; Sérgio Murilo Steffens: Visualization, Investigation ; Aldanéa Norma de Souza Silvestrin: Supervision, Investigation; Grace Serafim Claro: Visualization, Investigation; Hugo Alejandro Iskenderian: Visualization, Investigation; Bianca Hillmann: Visualization, Investigation, Reviewing; Liliam Gervasi: Investigation; Alberto Trapani Junior: Investigation; Patricia de Amorim Rodrigues: Investigation, Funding acquisition; Amanda de Souza Vieira: Investigation, Funding acquisition; Scheila Monteiro Evaristo Visualization, Investigation; Francisco Reis Tristão: Investigation, Funding acquisition; Fabiano da Silva Muniz: Investigation, Funding acquisition; Maria Veronica Nunes: Investigation, Funding acquisition; Nicole Zazula Beatriz: Investigation; Jhonathan Elpo: Investigation; Amanda Tiedje: Investigation; Louise Staudt Siqueira: Investigation; Marilin Sens: Investigation; Vitor Nandi: Investigation; Jessica Goedert Pereira: Investigation; Gustavo Costa Henrique: Investigation; Ana Paula Fritzen de Carvalho: Investigation; Ramon Carlos Pedroso de Morais: Investigation; Gustavo Giorgio de Cristo: Investigation; Maria Eduarda Hochsprung: Investigation; Ana Cristina Morais: Investigation; Rubens Centenaro: Investigation; Andrez Garcia Investigation; Bettina Heidenreich Silva: Investigation; Eluze Luz Ouriques: Investigation; Maria Eduarda Alves Ferreira: Investigation; Maria Eduarda Hames: Investigation; Maria Eduarda Paixão Gubert: Investigation; Milena Ronise Calegari: Investigation; Maria Luiza Baixo Martins: Investigation; Geovana Samuel Oliveira: Investigation; Marilia de Souza Marian: Investigation; Larissa Sell Sousa: Investigation; Marcelo da Silva Fedrizzi: Software, Validation.

All authors approved the final version of the manuscript and had access to the relevant study data and related analyses and vouch for the completeness and accuracy of the data presented.

## SUPPLEMENTARY TABLES

**Table S1.** Positive PCR results at more than 1 visit among MMRCoV clinical trial participants.

| Volunteer (allocation) | Group | Visit |
| --- | --- | --- |
| **Probable reinfection\*** |  |  |
| **023** | Vaccine | V1, V4 |
| **031** | Vaccine | V1, V4 |
| **048** | Placebo | V1, V6 |
| **052** | Vaccine | V1, V5 |
| **054** | Vaccine | V1, V5 |
| **204** | Vaccine | V2, V5 |
| **Persistent infection or reactivation**<br>**or SARS-CoV-2 RNA fragment<sup>†</sup>** |  |  |
| **008** | Vaccine | V5, V7 |
| **050** | Vaccine | V1, V2 |
| **095** | Placebo | V4, V5 |
| **118** | Vaccine | V5, V6, V7 |
| **170** | Placebo | V5, V7 |
| **220** | Placebo | V4, V5 |
| **346** | Vaccine | V3, V4 |
| **390** | Vaccine | V1, V3 |

\*Cases of probable reinfection were observed in participants who had a positive RT-PCR test using nasopharyngeal samples at least 3 months apart, interspersed with negative RT-PCR tests. The interval between positive RT-PCR tests ranged from 3 to 5 months. It was not possible to perform genetic sequencing of the samples.
†Positive RT-PCR tests at intervals of less than 3 months, in subsequent visits or interspersed with negative RT-PCR tests. These cases may be related to persistent viral infection, reactivation of infection or detection of SARS-CoV-2 RNA fragments.

**Table S2.** Local and systemic adverse events reported after injection of MMR vaccine or placebo in the MMRCoV clinical trial.

| Adverse events* | MMR Group<br>(N = 246)<br>N (%) | Placebo Group<br>(N = 178)<br>N (%) | p |
| --- | --- | --- | --- |
| **LOCAL** |  |  |  |
| Pain | 22 (8.9) | 8 (4.5) | 0.086 |
| Redness | 17 (6.9) | 2 (1.1) | 0.004 |
| Heat | 10 (4.1) | 1 (1.7) | 0.253 |
| Swelling | 11 (4.5) | 0 (0.0) | 0.003 |
| Other local symptoms<sup>†</sup> | 3 (1.2) | 0 (0.0) | 0.268 |
| **SYSTEMIC** |  |  |  |
| Loss of appetite | 4 (1.6) | 3 (1.7) | 1.000 |
| Headache | 46 (18.7) | 32 (18.0) | 0.899 |
| Muscle ache | 18 (7.3) | 9 (5.1) | 0.422 |
| Fever | 6 (2.4) | 2 (1.1) | 0.477 |
| Asthenia | 11 (4.5) | 9 (5.1) | 0.819 |
| Nausea | 6 (2.4) | 3 (1.7) | 0.740 |
| Other systemic symptoms<sup>‡</sup> | 27 (11.0) | 14 (7.9) | 0.321 |

\*The local and systemic adverse events were collected through a postvaccination diary, where the participant recorded all possible events up to 30 days after the injection. All reported adverse events occurred in the first week after injection and were considered mild.
<sup>†</sup>Other local symptoms reported by the participants were ecchymosis and pruritus.
<sup>‡</sup>Other systemic symptoms reported by the participants were joint pain, runny nose, diarrhoea, skin rash, cough, nasal congestion and dizziness.

